# The microbiological and physical properties of catheters for intermittent catheterization: A systematic review on the impact of reuse and cleaning methods

**DOI:** 10.1101/2021.04.28.21256230

**Authors:** Mark Grasdal, Matthias Walter, Andrei V. Krassioukov

## Abstract

This systematic review provides an up to date and comprehensive summary of the clinical evidence of the effectiveness of various cleaning methods of intermittent catheterization that have been proposed to prepare catheters for reuse. This systematic review is registered at PROSPERO (registration number: CRD42020176065). A key word search of Medline (OVID), Excerpta Medica dataBASE (EMBASE, OVID), Cumulative Index to Nursing and Allied Health Literature (CINAHL), Web of Science and the Cochrane Central Register of Controlled Trials (CENTRAL), in addition to manual searches of retrieved articles, was undertaken to identify all English, Russian and German language literature evaluating the effectiveness of various cleaning methods of intermittent catheterization. Studies selected for review included analytical experimental, prospective cohort, cross-sectional and case series study designs. Prospective cleaning methods analyzed included heat-based sterilization, chemical cleaning solutions, mechanical abrasion, photocatalytic sterilization, and combined methods. Studies that failed to assess the bacterial colonization or physical properties of catheters following cleaning were excluded. In total, 12 studies (i.e. 9 analytical experimental, 1 cohort study, 1 cross-sectional and 1 case series) were included. Two cleaning methods were identified as likely being most promising: five-minute submersion in 70% alcohol and the “Milton method”. Each eliminated bacterial colonization without affecting the physical properties of the catheters. All other cleaning methods were either non-bactericidal or caused gross visual or microscopic damage to the catheters, rendering their reuse unsafe. Additional higher-powered studies confirming the safety and efficacy of these cleaning methods must be obtained before we would feel comfortable challenging current clinical recommendations.

## Introduction

Intermittent catheterization (IC) is a method of urinary drainage commonly used by individuals with neurogenic lower urinary tract dysfunction (NLUTD)^1^, often as a result of a spinal cord injury (SCI). It is performed by inserting a catheter into the bladder via the urethra, allowing for the drainage of urine. Once the bladder is empty, the catheter is removed and discarded.

IC was traditionally performed using an aseptic technique^2^, however in 1972, Lapides et al. first demonstrated that using a clean technique, which they coined clean IC (CIC), was equally as effective at preventing urinary tract infections (UTIs).^3^ Thus, CIC is considered the “gold standard” for bladder emptying in individuals with NLUTD and sufficient manual dexterity.^4–6^

Presently, apart from only one manufacturer^7^ that we are aware of, the majority of manufactured catheters are clearly labeled as single-use devices. As such, these catheters are distributed without instructions for cleaning, although it is well known that due to their cost and limited availability in many countries, they are commonly reused.^7^ Catheter reuse is especially common in developing countries where it is not unheard of for some individuals to use the same catheter for greater than a year.^7^ Some consumers have stated that they would value having the option to reuse in concert with single-use if there was a safe and practical method of cleaning them following IC.^8^

In 2014 Prieto et al. suggested that the reuse of catheters was as safe as the single use of catheters^9^, although this work has since been retracted after an additional independent review of the data found crucial discrepancies of data extraction and analyses within the review.^10^ At the time of writing, the Center for Disease Control and Prevention (CDC) clinical recommendations^11^ state that further work must be done in this area before the reuse of catheters can be safely recommended. In Canada, the Canadian Urological Association (CUA)^12^ recommends that catheters should only be reused in specific clinical scenarios, and that users should be made aware that there is limited evidence to support the cleaning and safe reuse of single-use catheters.

There are many advantages to reusing catheters, namely convenience, reduced economic burden for consumers, and less environmental impact due to reduced waste.^13,14^ These advantages are offset by the burden of cleaning the catheters, cost of cleaning supplies, the concern that reusing catheters may cause the rate of UTIs to increase, and the added cost associated with a potential rise in lower urinary tract (LUT) complications.^8,13,15,16^ There is recent evidence to suggest that structural damage of the catheter itself from cleaning could put users at risk of contracting UTIs by their reuse due to an increased risk of urethral and bladder trauma.^17^ Therefore, it is important to understand how proposed methods of cleaning catheters for their reuse impacts the catheter’s structural integrity.

A variety of catheters have been marketed for IC over the years. Historically, the most commonly used catheters were constructed of materials including rubber, latex, silicone, polyvinyl chloride (PVC) and polyethylene.^18^ Today, PVC catheters are the most commonly used^19^, with some having a hydrophilic outer coating for ease of use. Still, because of the continued variability in materials used, cleaning methods that are effective for some catheters may not be suitable for others. To date, there is no reported consensus on the preferred method of cleaning, nor the period of time that a single catheter may be reused.^20^ Obtaining consensus is further complicated by the fact there is high variability and poor compliance when performing cleaning techniques, leading to an increased risk of bacterial colonization^20,21^ and LUT symptoms. Consequently, it is of great interest to determine if there are effective catheter cleaning methods to help ensure patient safety for those who choose to reuse them, and to potentially reduce the environmental impact and financial burden of single-use catheters onto the healthcare system.

To that end, this systematic review provides a comprehensive analysis of the effectiveness of various proposed methods of cleaning of catheters for IC to prepare them for their reuse. We were specifically interested in comparing the effectiveness of cleaning and reuse methods by evaluating the outcomes of bacterial colonization and change in physical properties. First, we examined the effectiveness of cleaning methods in terms of their ability to eliminate or diminish bacterial colonization; and second, we examine the impact of cleaning techniques on the physical properties of catheters.

## Methods

### Protocol and registration

The Preferred Reporting Items for Systematic Reviews and Meta-Analyses (PRISMA) guidelines were used to report our systematic review.^22^ A PROSPERO review protocol exists and can be accessed (registration number CRD42020176065).

### Eligibility criteria

We included all published studies thus far which investigate the properties of intermittent catheters and how they are affected by various cleaning procedures during re-use. We were specifically interested in studies focusing on either the relationship between cleaning during reuse and bacterial colonization of an intermittent catheter or the effects of cleaning and reuse on the physical properties of an intermittent catheter.

### Information sources and search strategy

A systematic review of all relevant literature, published from January 1, 1990 to May 20, 2020 was conducted using five databases (i.e., Medline [OVID], Excerpta Medica dataBASE [EMBASE, OVID], Cumulative Index to Nursing and Allied Health Literature [CINAHL], Web of Science and the Cochrane Central Register of Controlled Trials [CENTRAL]). The population key words were: “intermittent catheter”, “bladder catheter” and “urinary catheter”. These were paired with the terms: “clean”, “cleaning methods”, “cleaning solution”, “disinfect”, “decontaminate”, “sterilize” OR “physical”, “microscopic”, “mechanical” OR “reuse”. See Supplementary Appendix SA1 for the full search strategies used for each database.

### Study Selection

Studies were included for qualitative analysis if they met the following criteria: published in English, German or Russian language that evaluated the impact that various methods of cleaning catheters have on catheter bacterial colonization and changes in catheter physical properties. We included all study designs except reviews, book chapters, opinion papers, non-peer-reviewed work, conference abstracts or papers and studies where the full text was unavailable.

### Data extraction

Studies were collated and uploaded into Covidence.^23^ Independent reviewers (author 1 and 3) screened titles, abstracts, and full-texts; only eligible studies were included in the qualitative analysis. A third reviewer (author 2) resolved discrepancies. Figure 1 illustrates the PRISMA flow diagram. A consensus was achieved (between authors 1-3) on data to be extracted from studies, which included author and year of the study, study design, number of subjects and dropouts, inclusion/exclusion criteria, study aims, type of catheter, catheter inoculation method, catheter cleaning method, outcomes and conclusions, limitations and funding.

**Figure 1.**
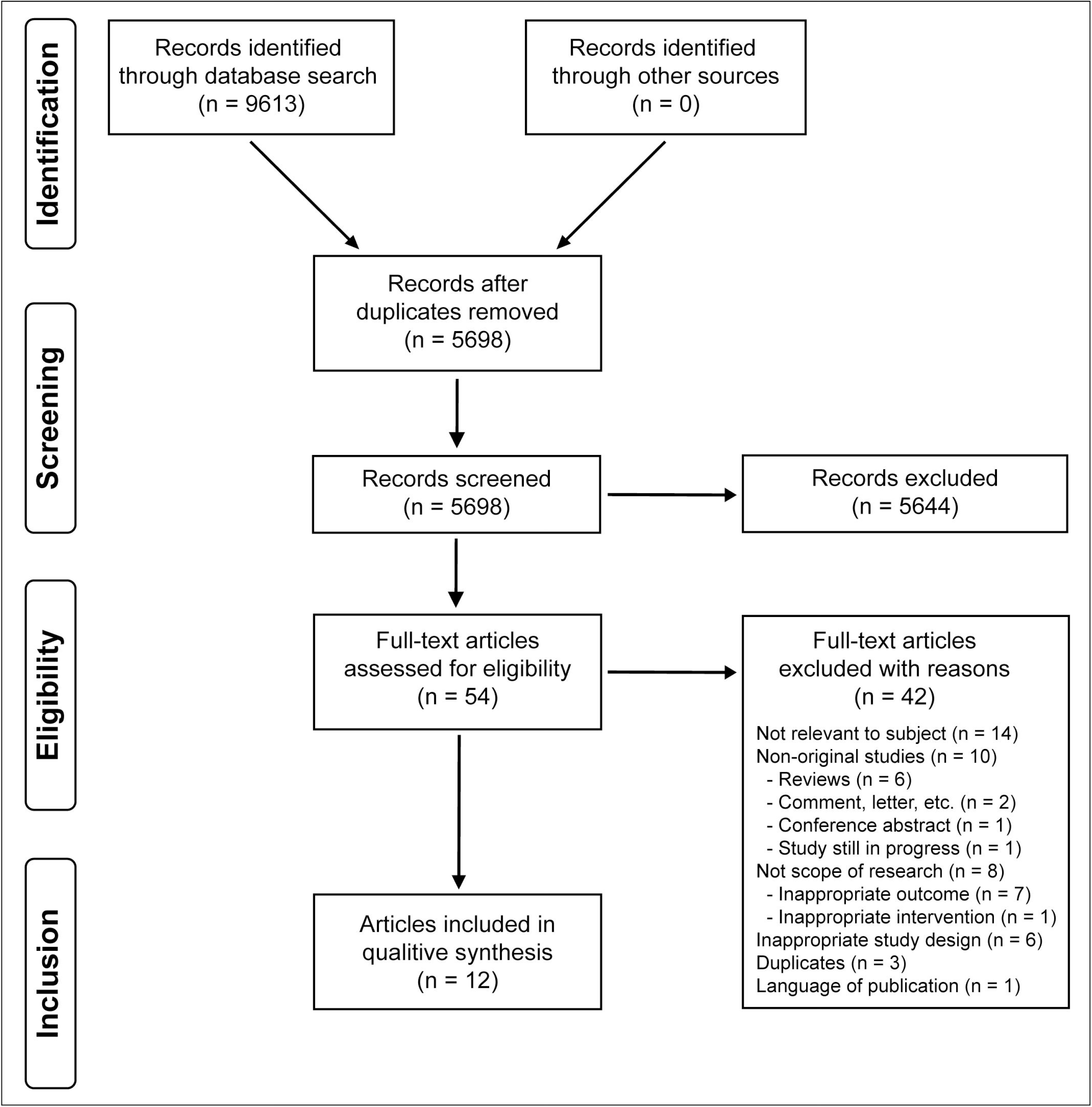
PRISMA flow diagram.

### Assessment of methodological quality

A quality assessment was completed for each study by two independent reviewers (author 1 and 3). Experimental studies were examined by the Joanna Briggs Institute Critical Appraisal Checklist for quasi-experimental studies^24^ to evaluate their methodological quality and therefore eligibility for inclusion. Cohort and cross-sectional studies were assessed according to the National Institute of Health quality assessment tool for observational cohort and cross-sectional studies.^25^ Case series studies were assessed according to the National Institute of Health quality assessment tool for case series studies.^26^ A third reviewer (author 2) resolved discrepancies.

### Risk of bias assessment

The risk of bias was assessed for eligible studies (i.e. cohort studies) in accordance with the Risk Of Bias in Non-randomized Studies - of Interventions (ROBINS-I).^27^

### Statistical analysis

Extracted data was organized in tabular form using Excel (Microsoft 365, version 2103). No meta-analysis or formal statistical tests for significance were performed.

## Results

The literature search yielded 9613 articles. After eliminating duplicates and reviewing the remaining titles and abstracts (Figure 1), a total of 12 studies^7,16,19,21,28–35^ that included outcomes of catheter bacterial colonization or changes in physical properties following cleaning of catheters for reuse were eligible and included (Table 1).

**Table 1.**
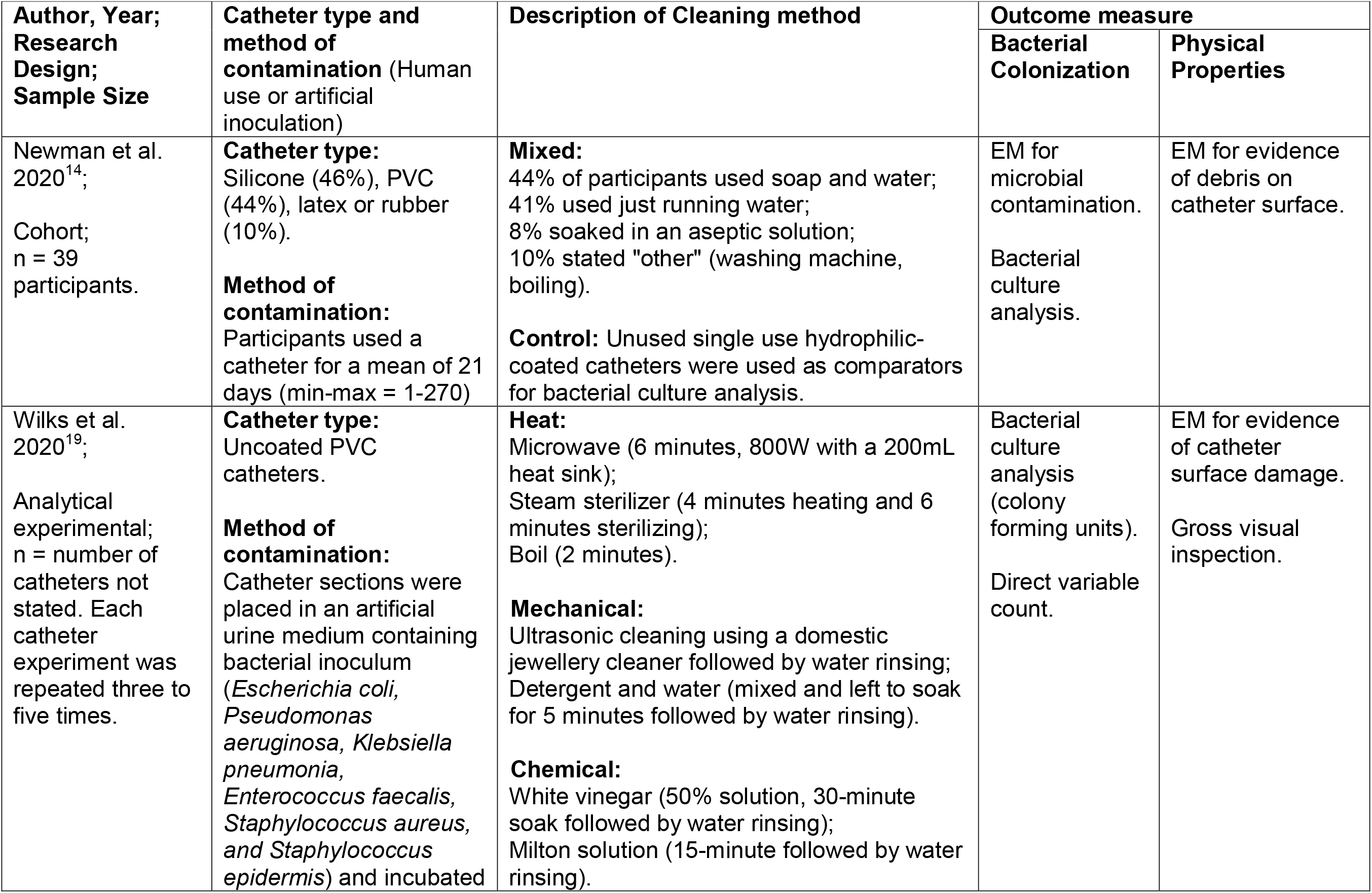
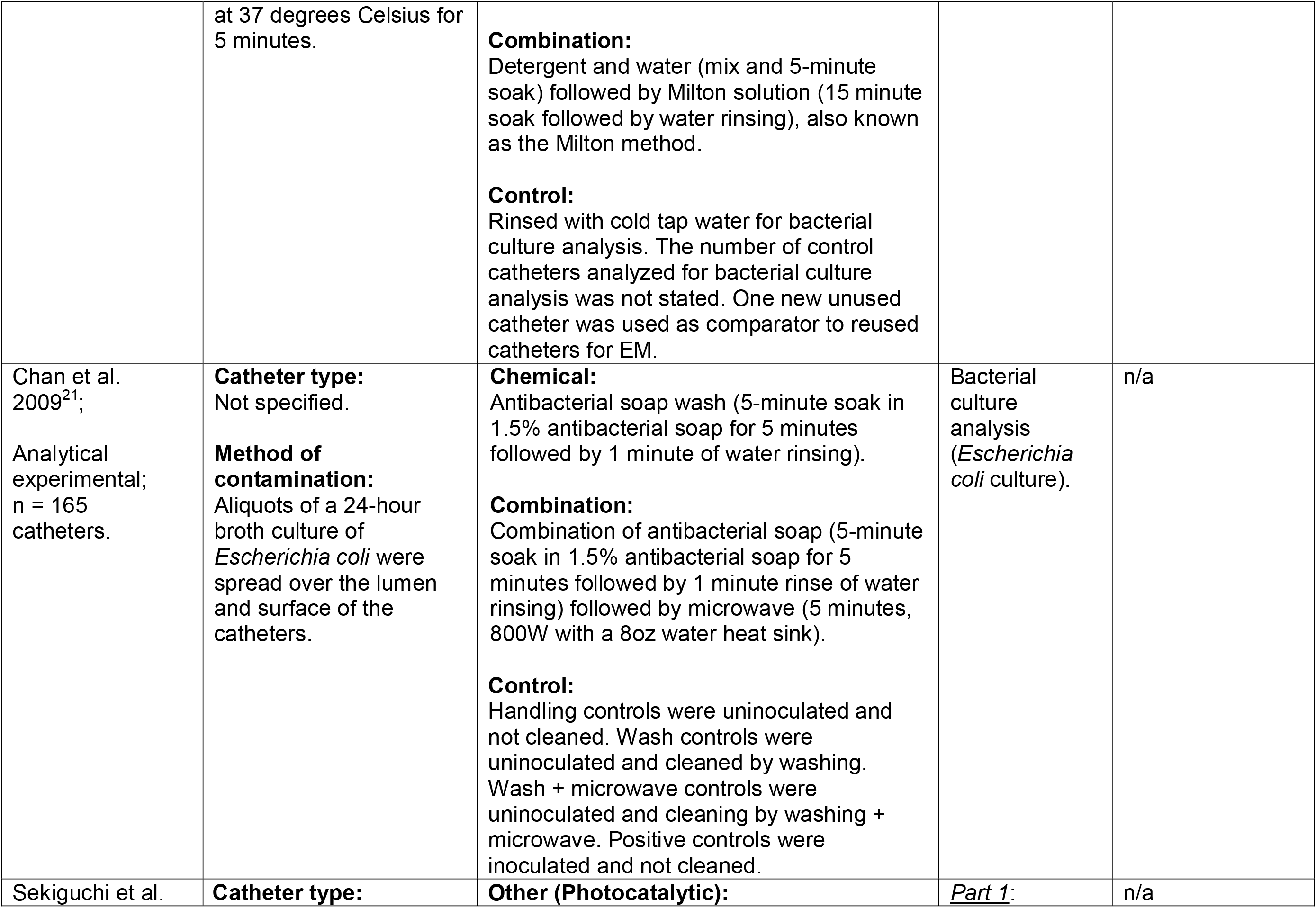
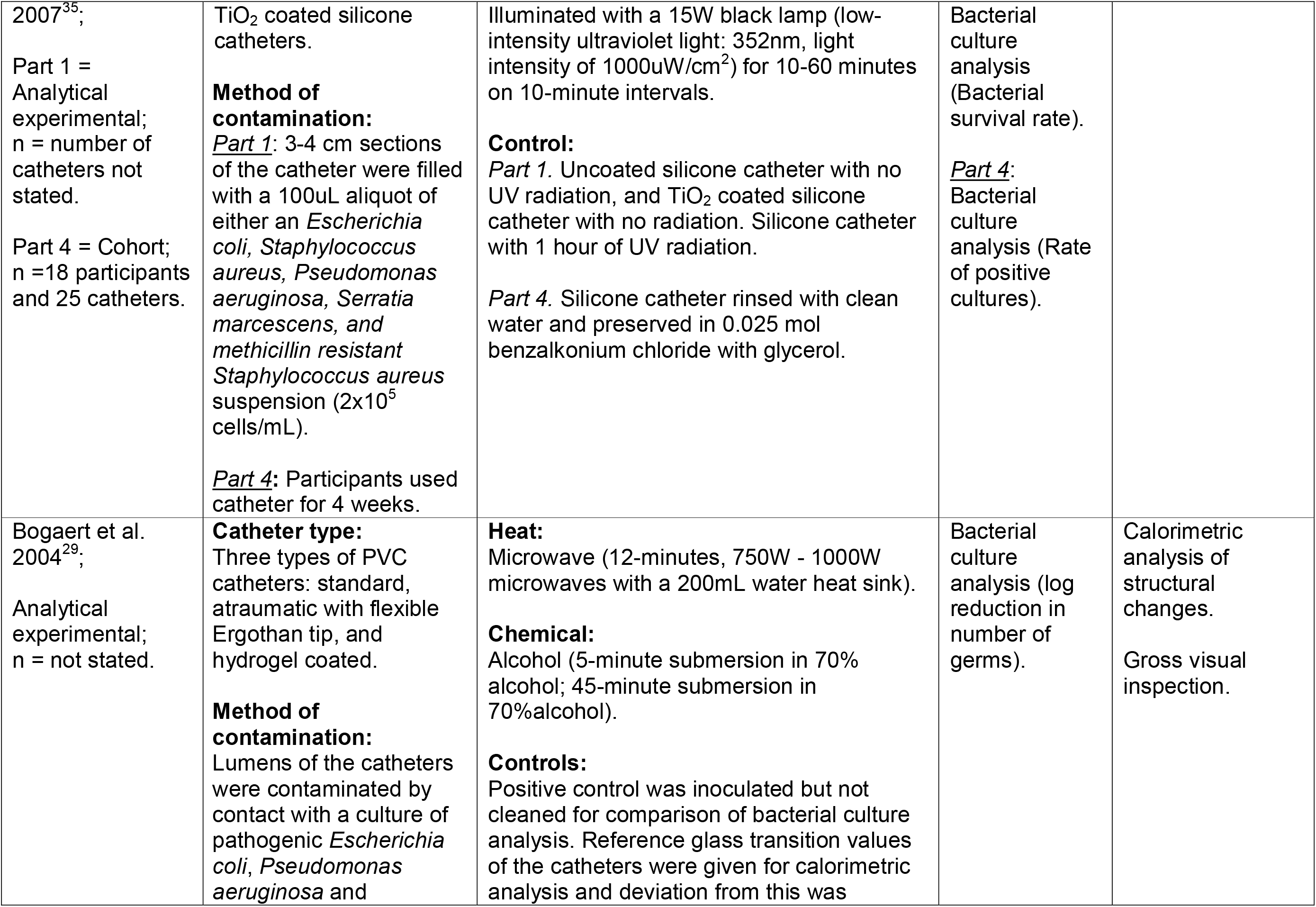
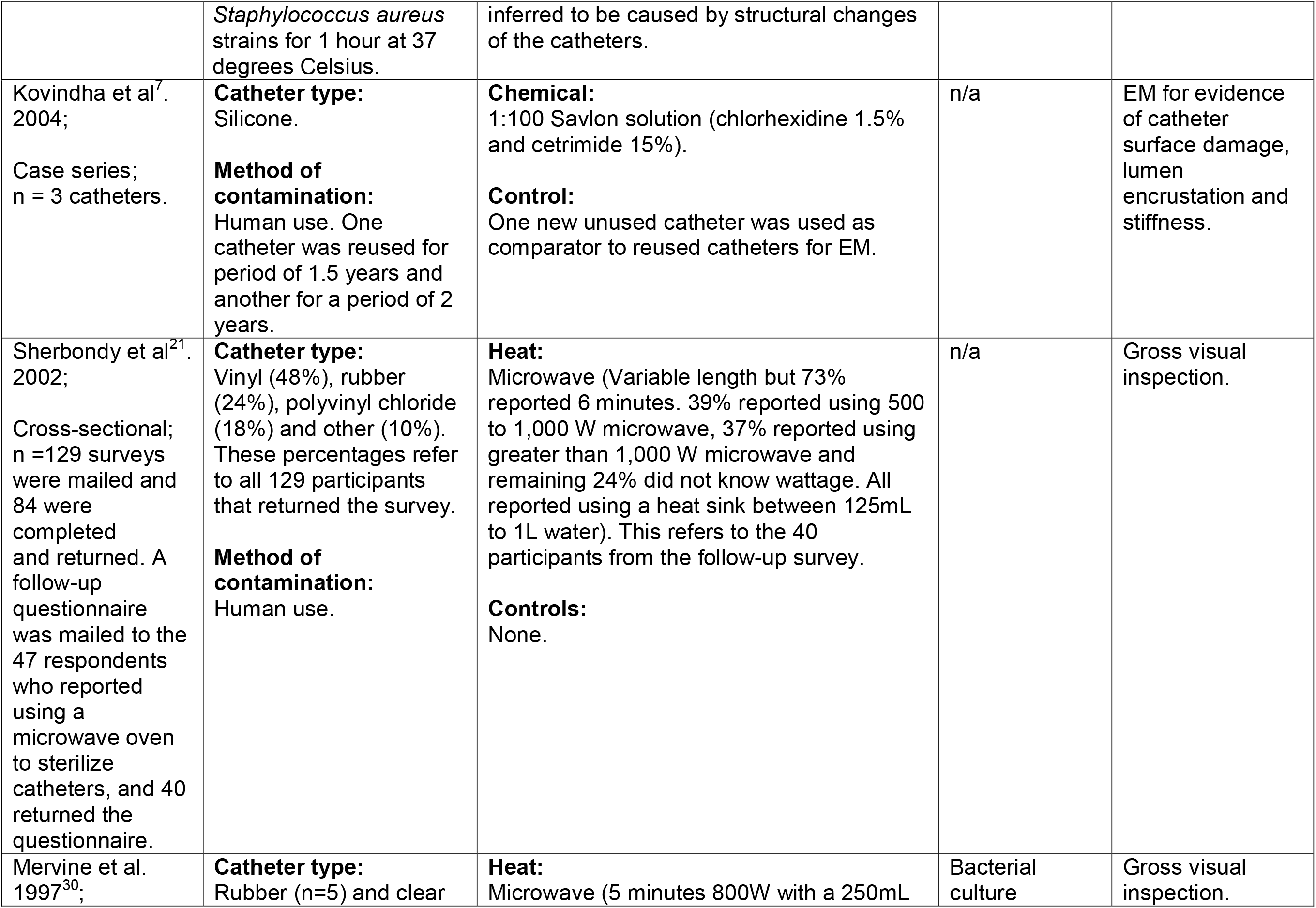
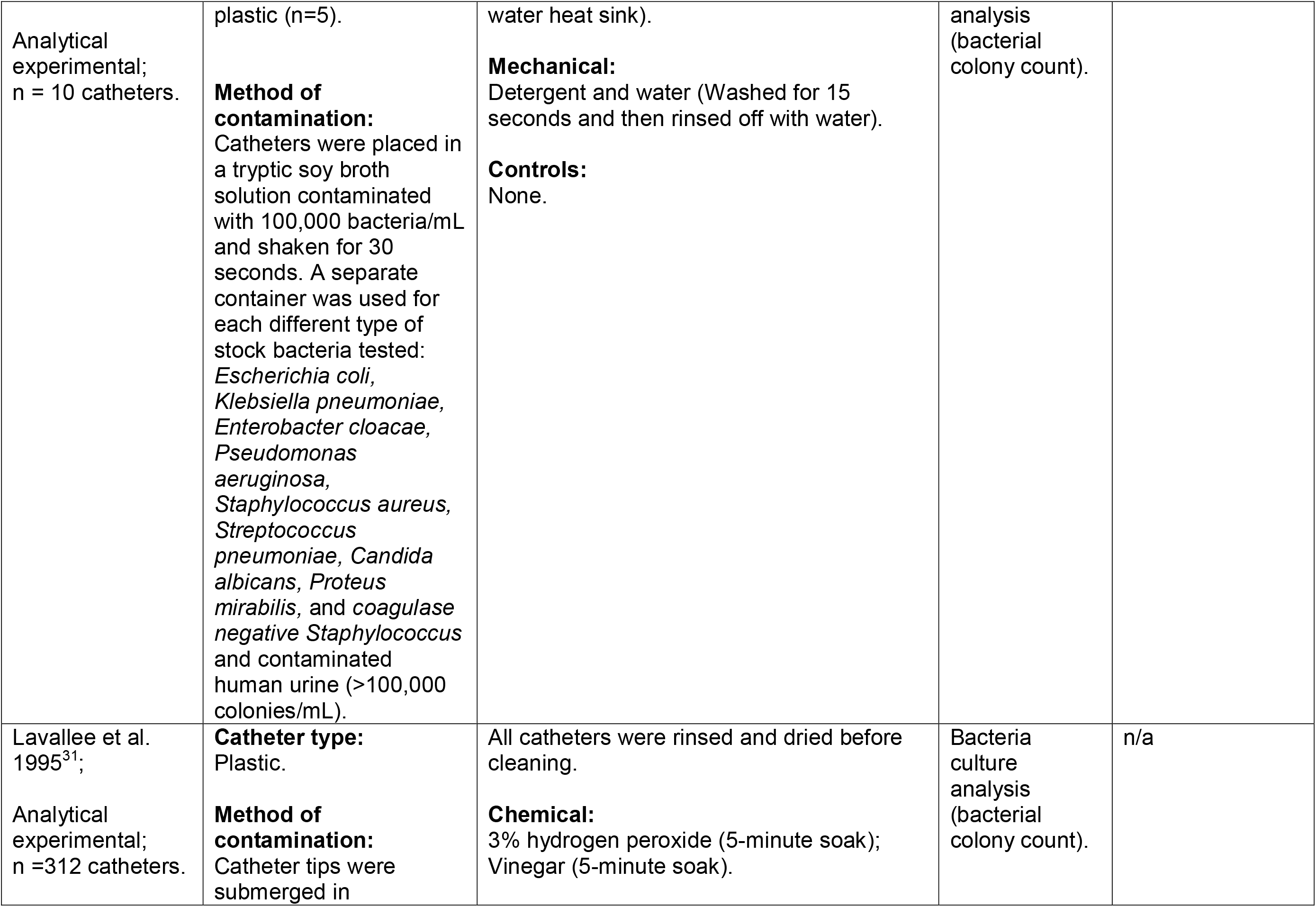
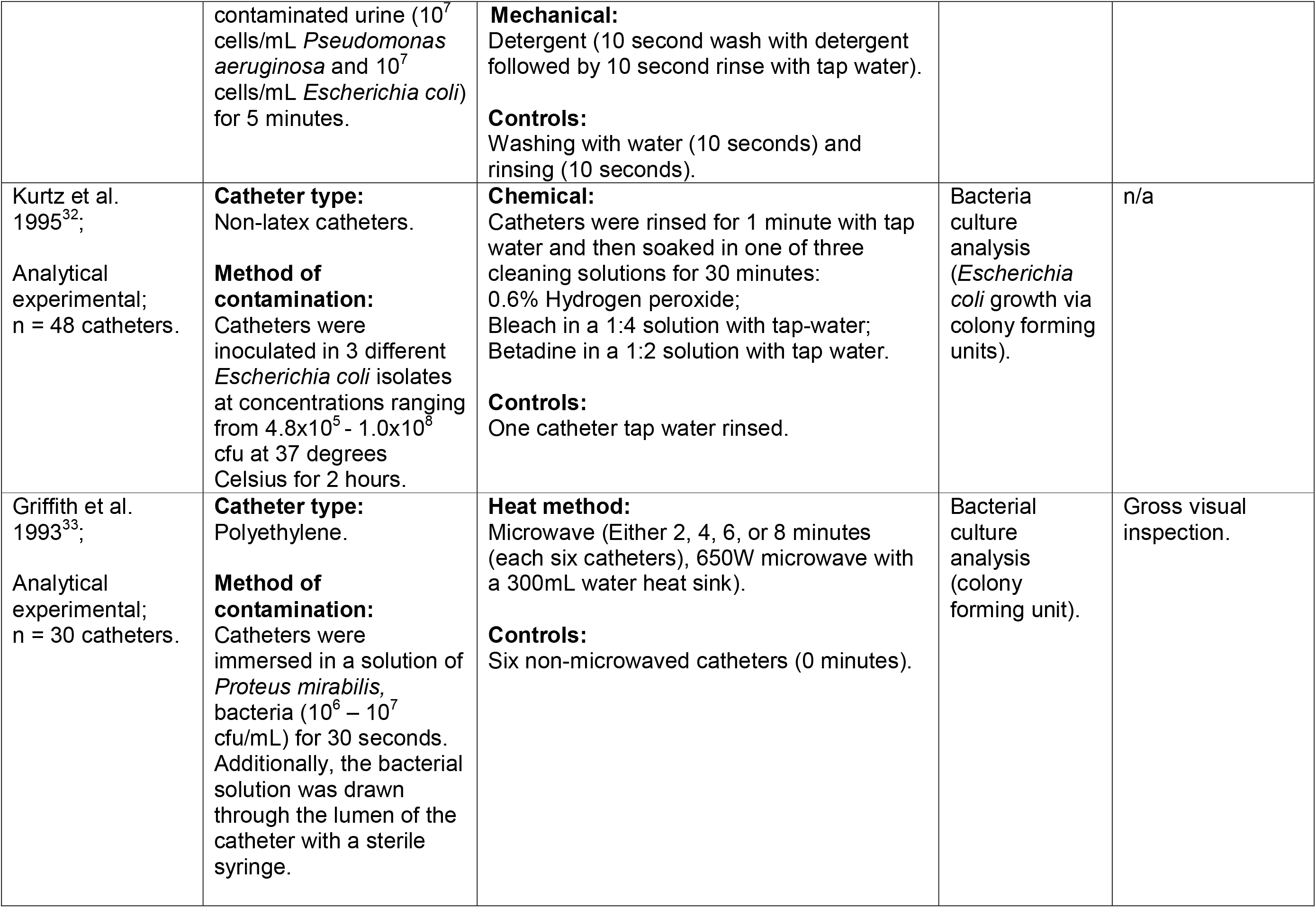
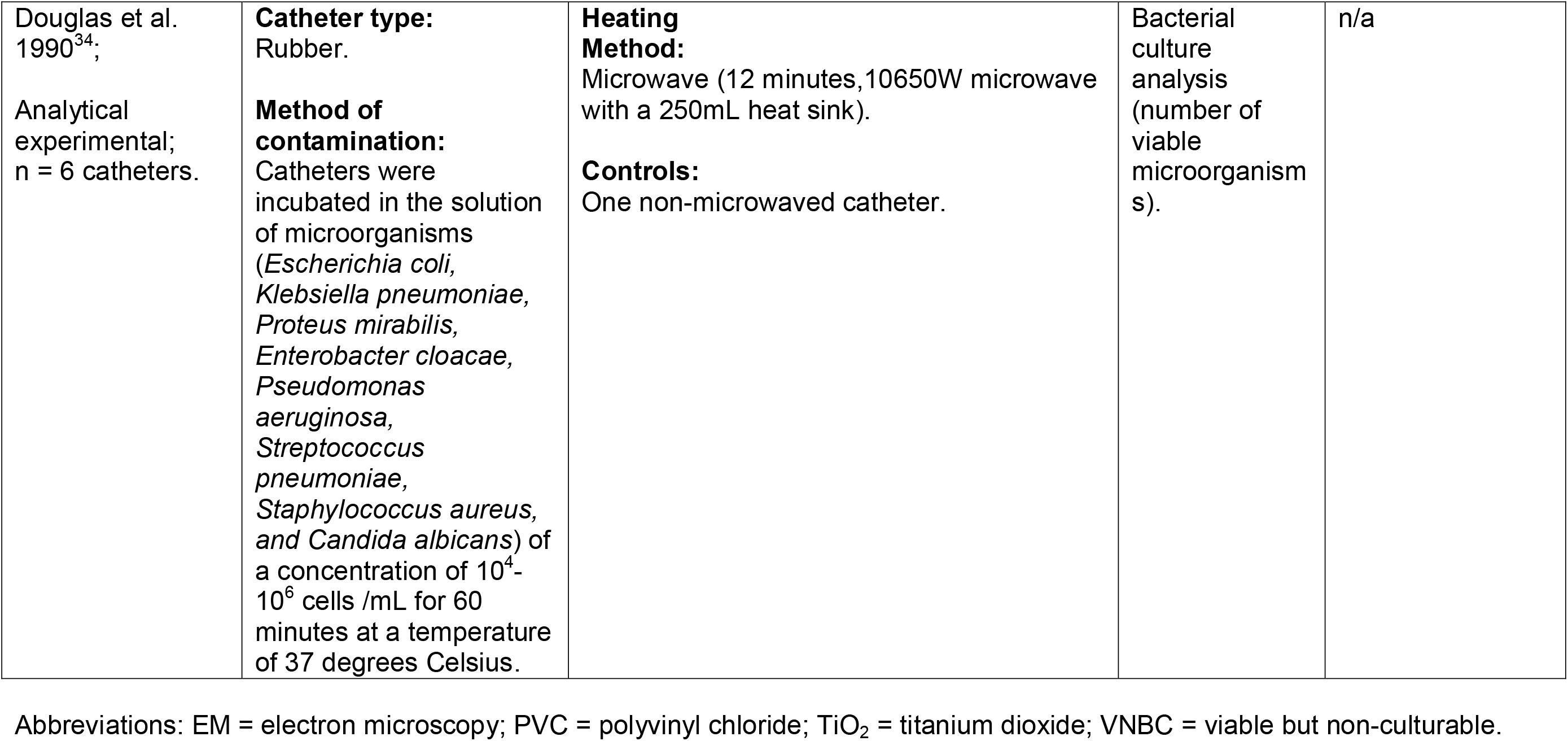
Overview of included studies.

### Description of the studies

Of the 12 studies included in this systematic review, nine were analytical experimental studies^19,28–35^, one of which was also partially a cohort study.^30^ The remaining were either a cohort^16^ or cross-sectional^21^ study and a case series.^7^ Based on presently published literature, multiple cleaning methods have been tested and proposed in order to decontaminate catheters including heat (microwave^19,21,29,31–33,35^, steam^19^, or boiling^19^), mechanical (ultrasonic^19^, detergent and rinsing^19,28,32,35^), chemical (bleach^34^, hydrogen peroxide^28,34^, betadine^34^, vinegar^19,28^, Milton solution^19^, Savlon solution^7^, 70% alcohol^31^), and other (photocatalytic^30^, mixture of methods^16^ or combination of two methods^19,35^). The majority of these studies (n = 10)^16,19,28–35^ used culture analysis as a microbiological technique to assess for the presence of bacteria. Several studies (n = 7)^7,16,19,21,29,31,32^ examined the impact of cleaning methods on physical properties of catheters, however only a minority (n = 4)^7,16,19,31^ examined the microscopic physical properties of the catheter. All 12 studies were grouped based on the type of cleaning methods analyzed, which included the following categories: heat-based, mechanical-based, chemical, combined and others. Next, the effectiveness of cleaning based on levels of bacterial colonization after using specific cleaning methods was determined (Tables 2-6). In the majority of studies, the bacterial colonization was either measured in terms of colony forming units (cfu) or log reduction depending on the quantification method reported. Changes in physical properties of catheters following various cleaning methods was then determined (Tables 2-6). These changes were evaluated macroscopically (i.e., gross visible inspection of changes and change in stiffness) and/or microscopically (by some form of Electron Microscopy (EM) or Differential Scanning Calorimetry (DSC)).

**Table 2.**
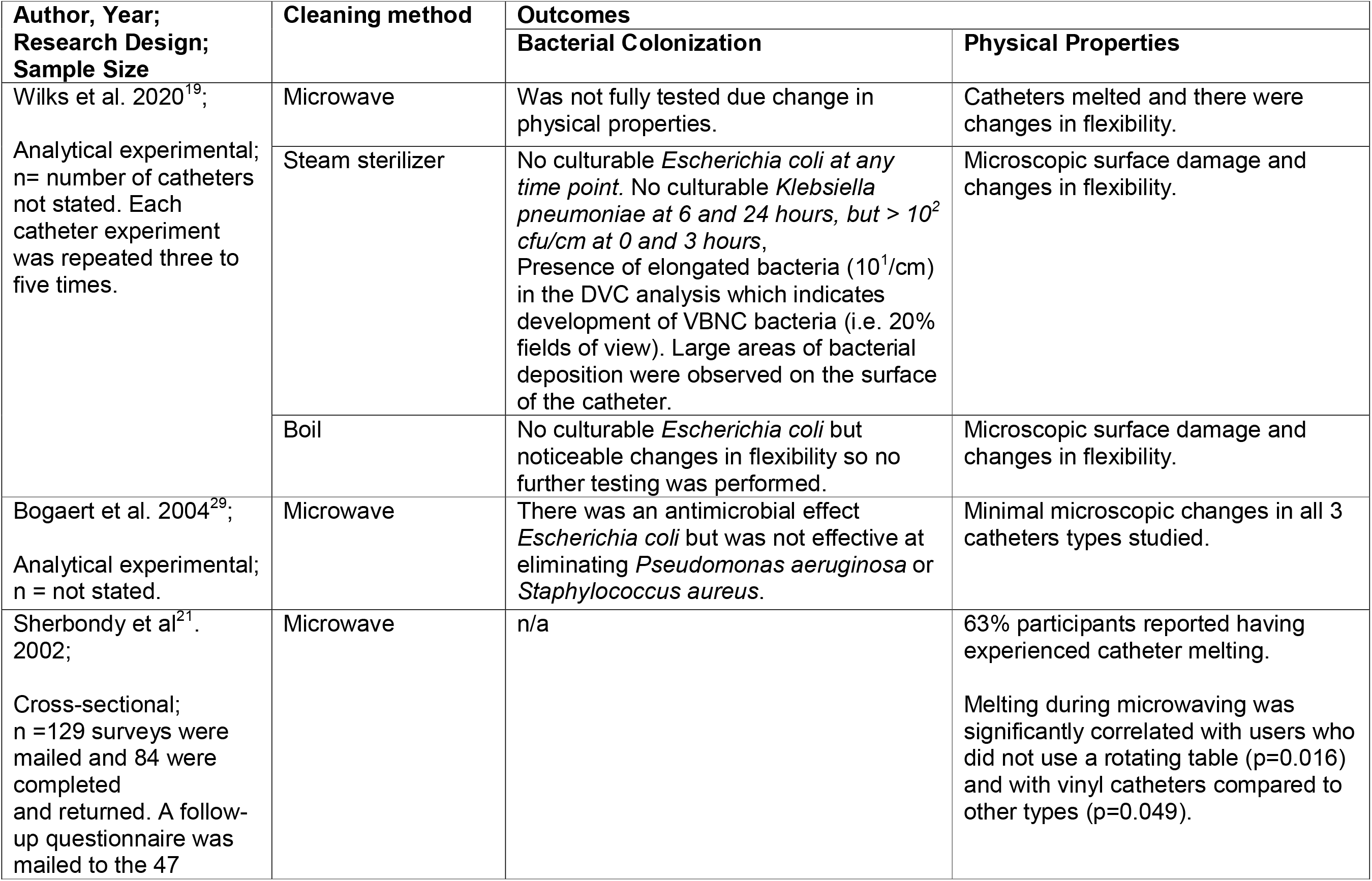
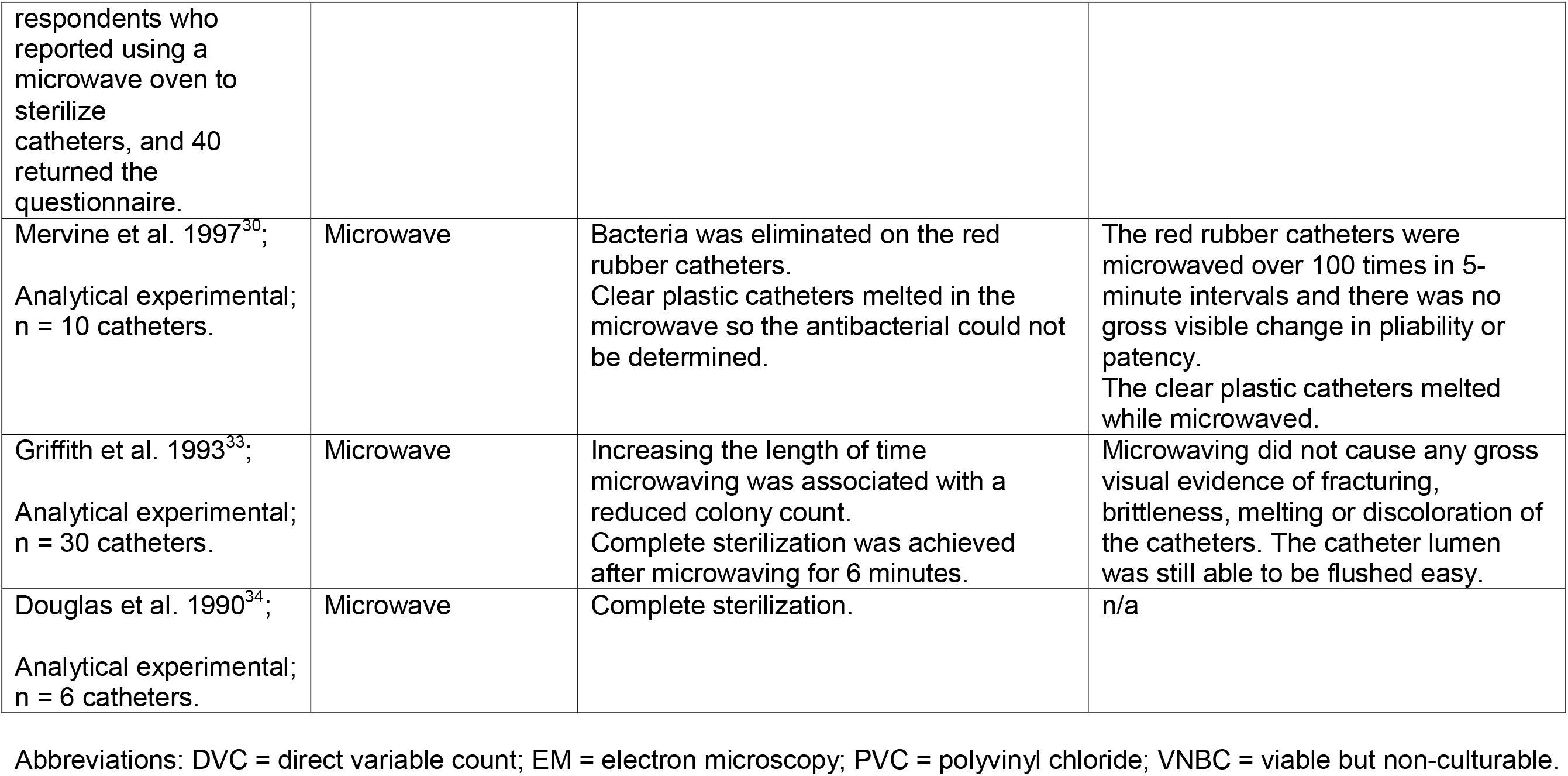
Outcomes of heat-based methods.

**Table 3.**
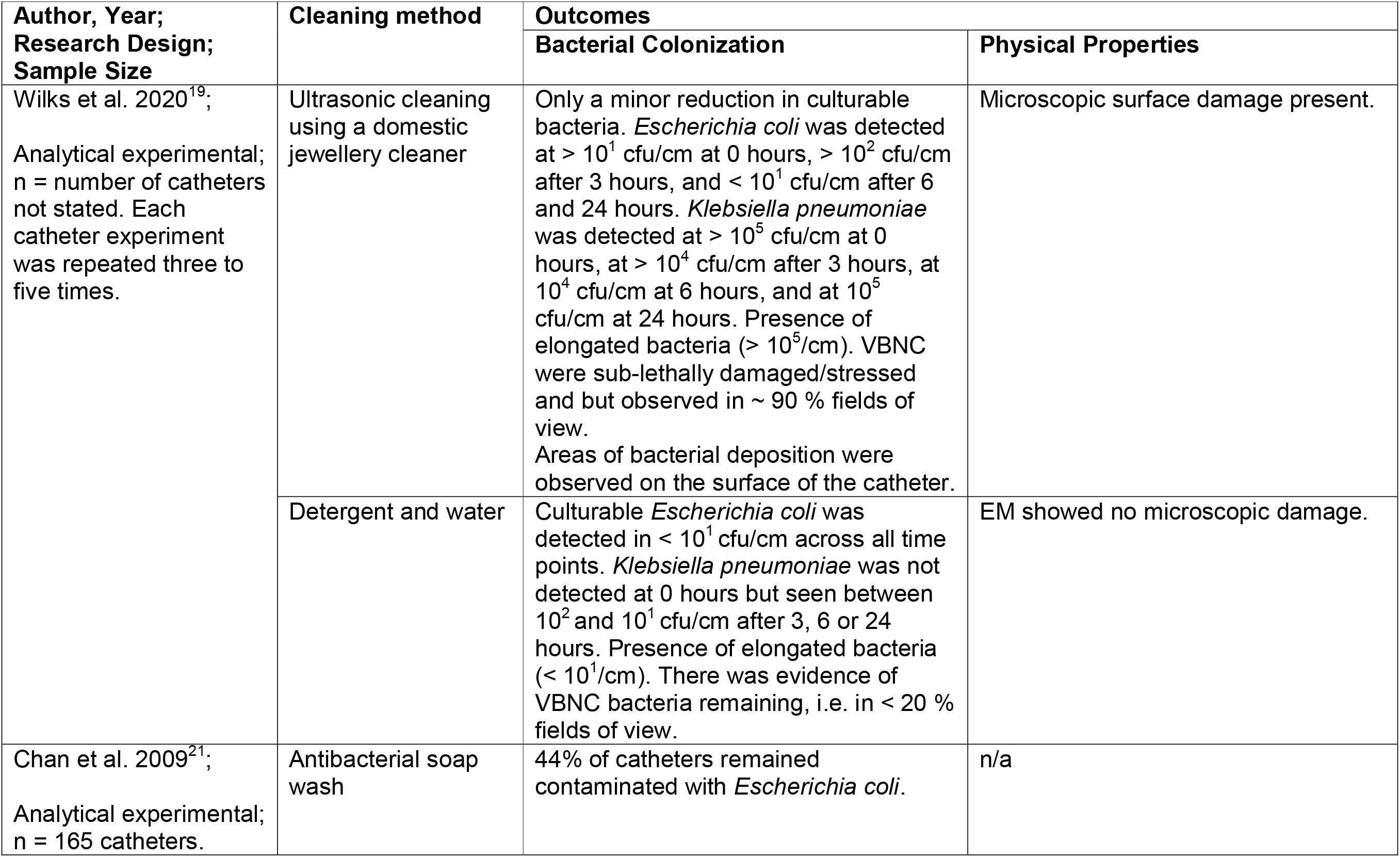
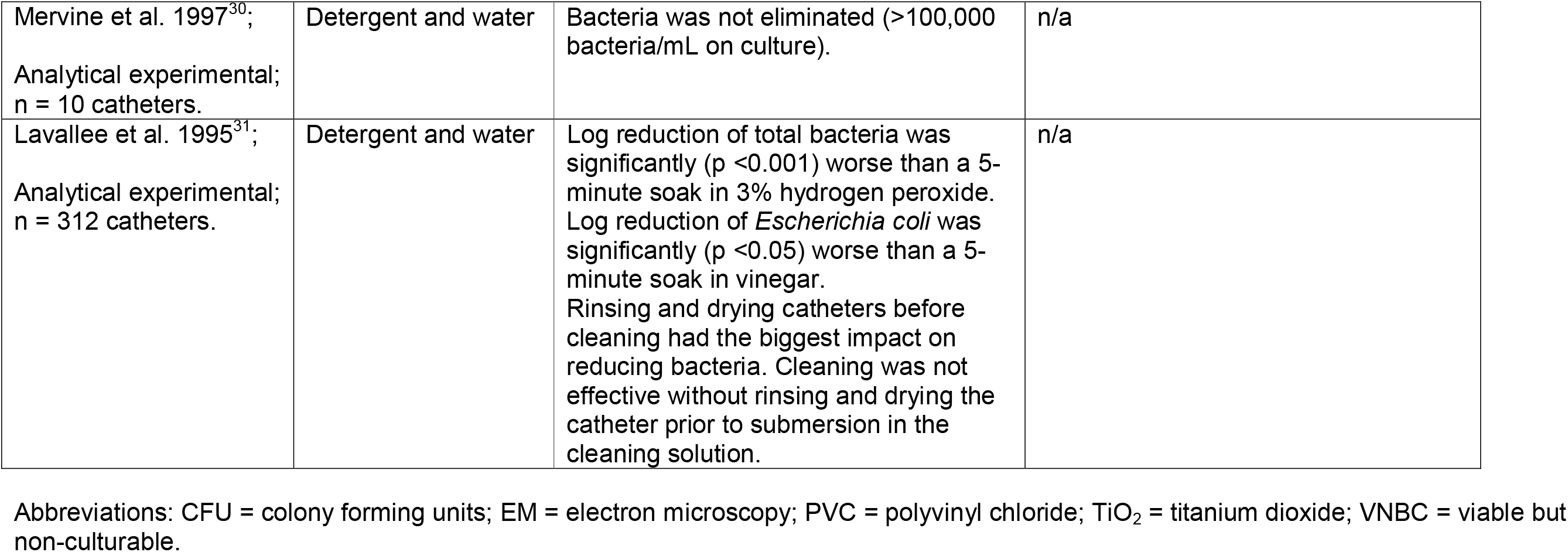
Outcomes of mechanical-based methods.

**Table 4.** Outcomes of chemical methods.

| Author, Year;<br>Research Design;<br>Sample Size | Cleaning method | Outcomes |  |
| --- | --- | --- | --- |
|  |  | Bacterial Colonization | Physical Properties |
| Wilks et al. 2020<sup>19</sup>;<br>Analytical<br>experimental;<br>n= number of<br>catheters not<br>stated. Each<br>catheter experiment<br>was repeated three<br>to five times. | Vinegar | Less than $10^1$ cfu/cm of culturable *Escherichia coli* was detected at 0, 3, 6 or 24 hours after exposure and there was no detectable culturable *Klebsiella pneumoniae* at all time point. Presence of elongated bacteria ($> 10^1$/cm). There was evidence of VBNC population remaining (i.e. ~ 20% fields of view). | No change. |
| | Milton solution | No culturable *Escherichia coli* detected at any time point. *Klebsiella pneumoniae* was detected at $10^3$ cfu/cm at 0 and 3 hours, $10^2$ cfu/cm at 6 hours but not culturable at 24 hours. When tested with full range of uropathogens, no culturable bacteria was observed after 24 hours of repeated exposure. Presence of elongated bacteria ($< 10^1$/cm). There was evidence of VBNC population remaining (i.e. $< 20\%$ fields of view). | EM showed no microscopic damage. |
| Bogaert et al. 2004<sup>29</sup>;<br>Analytical<br>experimental;<br>n = not stated. | 70% alcohol | Submersion for 5 minutes resulted in a complete antimicrobial effect on *Escherichia coli*, *Pseudomonas aeruginosa* and *Staphylococcus aureus* in all catheters.<br>There was no additional benefit to soaking the catheters for 45 minutes. | Soaking catheters for 5 minutes did not affect the physical properties of the catheters.<br>Soaking catheters for greater than 45 minutes in the 70% alcohol solution caused considerable microscopic changes in the physical properties of all catheters as demonstrated as increased glass transition values during calorimetric analysis. |
|  |  |  | It should be advised to not store catheters for reuse chronically in an alcohol solution as it causes significant structural damage of PVC catheters. |
| Kovindha et al.<sup>7</sup>.<br>2004;<br>Case series;<br>n=3 catheters. | Savlon solution | n/a | EM showed encrustation in lumen.<br>The reused catheters showed a 20% increase in stiffness compared to a new catheter. |
| Lavallee et al.<br>1995<sup>31</sup>;<br>Analytical<br>experimental;<br>n = 312 catheters. | 3% hydrogen peroxide | Log reduction of total bacteria was significantly (p <0.001) better than washing with detergent and water.<br>Log reduction of *Escherichia coli* was significantly (p <0.05) better than using vinegar.<br>Log reduction of *Pseudomonas aeruginosa* was significantly (p <0.05) worse than using vinegar.<br>Rinsing and drying catheters before cleaning had the biggest impact on reducing bacteria.<br>Cleaning was not effective without rinsing and drying the catheter prior to submersion in the cleaning solution. | n/a |
|  | Vinegar | Log reduction of *Escherichia coli* was significantly (p <0.05) worse than using 3% hydrogen peroxide.<br>Log reduction of *Pseudomonas aeruginosa* was significantly (p <0.05) better than using 3% hydrogen peroxide.<br>Rinsing and drying catheters before cleaning had the biggest impact on reducing bacteria.<br>Cleaning was not effective without rinsing and drying the catheter prior to submersion in the cleaning solution. |  |
| Kurtz et al. 1995<sup>32</sup>;<br>Analytical<br>experimental;<br>n = 48 catheters. | 0.6% Hydrogen<br>peroxide | All three procedures were equal and effective,<br>completely preventing *Escherichia coli* growth<br>when measured 48 hours post cleaning. | n/a |
|  | Bleach solution |  |  |
|  | Betadine solution |  |  |

Abbreviations: EM = electron microscopy; PVC = polyvinyl chloride; VNBC = viable but nonculturable; TiO<sub>2</sub> = titanium dioxide

**Table 5.** Outcomes of combined, mixed and other methods.

| Author, Year;<br>Research<br>Design;<br>Sample Size | Cleaning method | Outcomes |  |
| --- | --- | --- | --- |
|  |  | Bacterial Colonization | Physical Properties |
| Newman et al,<br>2020<sup>14</sup>;<br>Cohort;<br>n = 39<br>participants. | Mixed | 74% of reused catheters had microbial contamination: 20 different bacterial species were found in total. A biofilm was found in 20% of the reused catheters. Viable microorganisms could be cultured in 67% of the samples. | Debris contamination was found on all reused catheters. EM showed the debris to be a mixture of residuals from urine, lubricant, soap, detergents and cells. |
| Wilks et al.,<br>2020<sup>19</sup>;<br>Analytical<br>experimental;<br>n = number of<br>catheters not<br>stated. Each<br>catheter<br>experiment was<br>repeated three to<br>five times. | Combined: Detergent<br>and water + Milton<br>solution also known as<br>"Milton method" | Total bactericidal effect on all tested uropathogens, hence no elongated bacteria per 1cm catheter section and 0 % fields of view with elongated bacteria. | No change. |
| Chan et al.<br>2009<sup>21</sup>;<br>Analytical<br>experimental;<br>n = 165<br>catheters. | Combined:<br>Antibacterial wash +<br>microwave | 26% of catheters washed with antibacterial soap remained contaminated with *Escherichia coli* | n/a |
| Sekiguchi et al.<br>2007<sup>35</sup>; | Photocatalytic | <u>Part 1</u>: The photocatalytic effect of the TiO<sub>2</sub> catheters reduced the survival rate of all bacteria to a negligible level within | n/a |
| Part 1 =<br>Analytical<br>experimental,<br>n = number of<br>catheters not<br>stated.<br>Part 4 = Cohort;<br>n =18 participants<br>and 25 catheters. |  | 60 minutes of black light illumination. The<br>antibacterial effect was stable after 200<br>uses.<br><u>Part 4:</u> 20% positive culture rate. |  |

Abbreviations: EM = electron microscopy; PVC = polyvinyl chloride; VNBC = viable but non-culturable; TiO<sub>2</sub> = titanium dioxide

**Table 6.** Outcomes of control methods.

| Author, Year;<br>Research Design;<br>Sample Size | Outcome |  |
| --- | --- | --- |
|  | Bacterial Colonization | Physical Properties |
| Newman et al. 2020<sup>14</sup>;<br>Cohort;<br>n = 39 participants. | Unused catheter had no bacterial contamination. | No EM control catheter. |
| Wilks et al. 2020<sup>19</sup>;<br>Analytical experimental;<br>n = number of catheters not stated. Each catheter experiment was repeated three to five times. | Consistently high values of culturable bacteria as well as VNCB present following tap water rinse. | Highly disordered but no damage or evidence of bacterial attachment. |
| Chan et al. 2009<sup>21</sup>;<br>Analytical experimental;<br>n = 165 catheters. | Handling and washing controls were negative for growth of the indicator organisms, but had growth from contaminants which the authors concluded were from the packaging materials.<br>89% of positive controls had *Escherichia coli* growth at 7 days. | n/a |
| Sekiguchi et al. 2007<sup>35</sup>;<br>Part 1 = Analytical experimental;<br>n = number of catheters not stated.<br>Part 4 = Cohort;<br>n = 18 participants and 25 catheters. | *Part 1.* Inoculated Silicone and TiO<sub>2</sub> coated silicone catheters that were not exposed to UV radiation had 70% and 80% *Escherichia coli* survival rate after 1 hour. Silicone catheter exposed to 1 hour of UV radiation had about 50% survival rate of *Escherichia coli* after 1 hour.<br>*Part 4.* 60% (6/10) catheters had positive cultures. | n/a |
| Bogaert et al. 2004<sup>29</sup>; | Inoculated uncleaned catheters had at | n/a |
| Analytical experimental;<br>n = not stated. | minimum 2 log greater of *Escherichia coli*,<br>*Pseudomonas aeruginosa* and<br>*Staphylococcus aureus* in comparison to<br>microwave and alcohol cleaned catheters. |  |
| Kovindha et al.<sup>7</sup>. 2004;<br>Case series;<br>n = 3 catheters. | n/a | Unused catheter had no evidence of<br>surface damage. |
| Lavallee et al. 1995<sup>31</sup>;<br>Analytical experimental;<br>n = 312 catheters. | Significantly higher amounts of<br>*Pseudomonas aeruginosa* and<br>*Escherichia coli* then peroxide, vinegar.<br>There was no significant difference<br>between the water rinsing control and<br>detergent rinse. | n/a |
| Kurtz et al. 1995<sup>32</sup>;<br>Analytical experimental;<br>n = 48 catheters. | Positive cultures in tap water rinse control<br>($5.2 \times 10^2$ cfu/mL). | n/a |
| Griffith et al. 1993<sup>33</sup>;<br>Analytical experimental;<br>n = 30 catheters. | The non-microwaved control catheter<br>produced a mean of $7.8 \times 10^4$ cfu per<br>catheter. | n/a |
| Douglas et al. 1990<sup>34</sup>;<br>Analytical experimental;<br>n = 6 catheters. | Control catheters had $10^4$-$10^6$ organisms<br>per catheter. | n/a |

Abbreviations: EM = electron microscopy; PVC = polyvinyl chloride; VNBC = viable but non-culturable; TiO<sub>2</sub> = titanium dioxid

### Quality of evidence and risk of bias

Overall, quality was fair for all included studies. See Supplementary Appendix SA2 for full details of quality assessment. The risk of bias was assessed in accordance with ROBINS-I for all eligible studies, i.e. the two cohort studies (see Figure 2). Risk-of-bias assessment showed either serious or critical risk of bias. All other studies did not qualify for risk of bias analysis since only catheters and not patients were reported.

**Figure 2.**
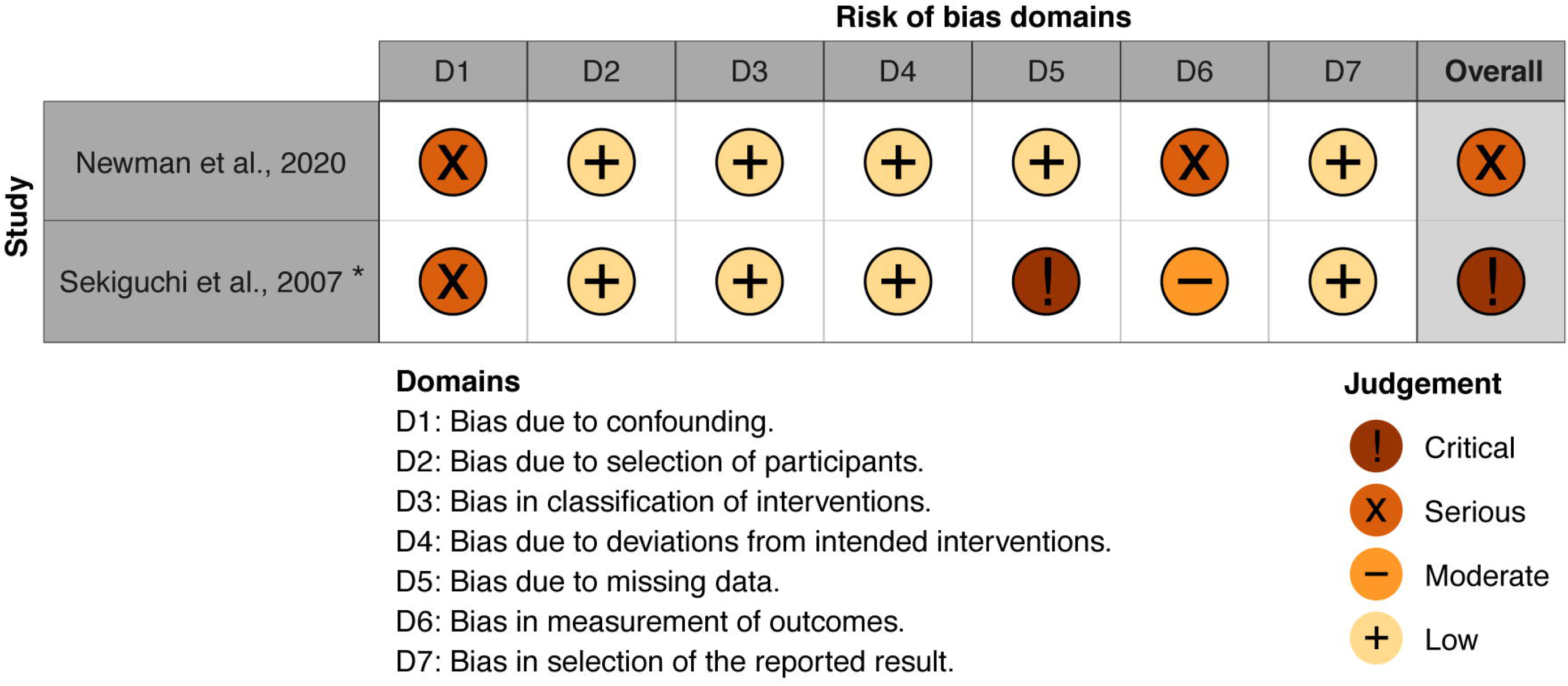
Risk of bias. * This publication consists of a multiple parts, i.e. analytical experimental part and cohort. The risk of bias analysis only addresses the cohort part of this publication.

### Heat-based methods

Among heat-based cleaning methods the microwave treatment (n = 6) ^19,21,29,31–33^ was the most commonly used followed by boiling (n = 1)^19^ and steam sterilization (n = 1).^19^ Overall, it was generally agreed that upon increasing the length of microwaving time resulted in a reduction in colony counts. Rubber catheters were reportedly able to be sterilized by microwave treatment^29,32,33^, but none of these studies fully analyzed the impact on the structural integrity of the catheters. When analyzing non-rubber catheters, none of the microwave treatments studied were as effective at reducing bacterial colony counts of *Escherichia (E*.*) coli, Pseudomonas (P*.*) aeruginosa* and *Staphylococcus (S*.*) aureus*, while also maintaining catheter structural integrity as were the chemical and combined methods (e.g. alcohol, “Milton method”)^19,31^. In two studies^19,32^, investigators were not able to obtain bacterial colonization data as the physical properties of the catheters changed to dramatically for the study to continue. Boiling in tap water for two minutes resulted in undetectable levels of *E. coli* but also led to the PVC catheter being damaged. Other uropathogens were not tested in this study due to evidence of catheter damage.^19^ Steam cleaning also resulted in undetectable *E. coli* or *Klebsiella (K*.*) pneumoniae*. However, there was evidence of some viable but non culturable bacterial population (VNCB).^19^

### Mechanical-based methods

The use of detergent and water was the most common mechanical cleaning method (n = 4)^19,28,32,35^, followed by ultrasonic jewelry cleaner (n = 1).^19^ Although these methods have demonstrated some reduction in bacterial counts, neither of these methods were effective for sterilisation. For example, Chan et al. showed that 44% of PVC catheters still had a positive culture following soaking in a 1.5% solution of antibacterial soap for 5 minutes followed by a 1-minute water wash.^35^ Wilks et al. reported that the use of a domestic jewelry cleaner was also not an effective cleaning method as the action of the cleaner was not bactericidal and damaged the catheter surface.^19^

### Chemical methods

There were a variety of cleaning solutions examined (n = 9).^7,19,28,31,34^ Submerging PVC catheters in a 70% alcohol solution for 5 minutes (n = 1)^31^ was the most effective as there was both bacterial sterilization (6-log colony count reduction) and showed no catheter damage. Another study determined that soaking plastic catheters for 5 minutes in 3% peroxide solution (n = 1)^28^ was not fully bactericidal but reduced the bacterial colony count by nearly 3 log. Similarly, it was found that soaking non-latex catheters in 0.6% hydrogen peroxide (n = 1), 1:4 solution of bleach in tap water (n = 1), or a 1:2 solution of betadine in tap water (n = 1) for 30 minutes were all effective at preventing *E*.*coli* growth 48 hours post cleaning, but there was no analysis for structural damage.^34^ Soaking in vinegar (n = 2)^19,28^ was not effective at eliminating most uropathogens, although it did not affect the physical properties of the catheters. Additionally, it was found that the use of Milton solution (diluted Milton concentrate with tap water as per manufacturer’s instructions, 0.6% sodium hypochlorite final concentration) (n = 1)^19^ resulted in no detectable culturable bacteria after repeated contamination and decontamination events over 24 hours when tested with a full range of uropathogens; however, there was evidence of some VNCB population remaining. A cross-sectional study by Kovindha et al. followed participants who kept their reusable silicone catheters in a Savlon solution between uses over the course of 1-2 years. These reused catheters showed encrustation of the lumen and an increase in stiffness of 20% relative to a new catheter.^7^ Finally, it should be noted that in general cleaning was not as effective without rinsing and drying the catheter prior to submersion in the cleaning solution.^28^

### Combined, mixed and other methods

There were two combined methods reported. The first combined an antibacterial soap wash for 5 minutes followed by microwaving in an 800W microwave for 5 minutes (n = 1).^35^ This was found to be more effective than washing with antibacterial soap alone, but still found that 24% of catheters had a positive *E. coli* culture. The other, coined the “Milton method” by Wilks et al, was found to be more effective and was performed by soaking a catheter in hot soapy water for 5 minutes, rinsed with water, left to soak in Milton solution for 15 minutes and then rinsed off with tap water again (n = 1).^19^ This resulted in undetectable *E. coli* levels, no evidence of a VNCB population and showed no structure damage to the uncoated PVC catheters. Sekiguchi et al. have developed a titanium dioxide (TiO_2_) coated silicone catheter which has a photocatalytic antimicrobial effect using only UVA illumination.^30^ After 60 minutes of black light illumination using a 15W 352nm black lamp with intensity of 1000uW/cm^2^, it was demonstrated that the photocatalytic effect of the TiO_2_ catheters reduced the survival of all bacteria tested (*Serratia marcescens, methicillin-sensitive S. aureus, methicillin-resistant S. aureus, P. aeruginosa*) to a negligible level. These catheters reportedly maintained their antibacterial effect after 200 uses. There was no data collected about how the sterilization affected the structure of the catheters. Another recent study by Newman et al. did not control for cleaning methods (participants choice and a mix of methods were reportedly used) but found that after a mean reuse of 21 days, 74% of catheters had microbial contamination and 20 different bacterial species were found in total. A biofilm was found in 20% of the reused catheters and viable microorganisms could be cultured in 67% of the samples. There was debris contamination found on all catheters which EM showed to be a mixture of residuals from urine, lubricant, soap, detergents and cells.^16^

### Controls

Unused catheters were found to have no microbial contamination (n=2)^16,35^, except in one study where growth of contaminants was found due to the packaging materials of the catheters.^35^ Likewise unused catheters had no surface damage as determined by EM (n=2).^7,19^ Catheters that were inoculated and not cleaned, or inoculated and rinsed with tap water only, had consistently higher bacterial colonization than catheters that had been cleaned by heat, mechanical, chemical, combined, or other methods (n=7).^19,29–31,33–35^ In a cohort trial by Sekiguchi et al. the control silicone catheters (n=10), which were rinsed with clean water and preserved in 0.025 mol benzalkonium chloride with glycerol, had a 60% positive culture rate versus a 20% positive culture rate from the TiO_2_ coated catheters (n=15) which were sterilized by the photocatalytic effect.^30^

## Discussion

This systematic review provides an up to date and comprehensive summary of the clinical evidence of the effectiveness of various cleaning methods of IC that have been proposed to prepare catheters for reuse. This analysis will help guide future clinical recommendations and is applicable to both health care providers and individuals requiring the use of catheters for IC.

The available evidence suggests that the effectiveness of microwave sterilization greatly depends on the material of the catheter. Rubber catheters seem to be able to withstand repeated microwave sterilizations with a common household microwave without incurring macroscopic changes.^29,32,33^ It should be noted that to date, no assessment of microscopic structural damage has been published after microwaving rubber catheters. Reported time to sterilize seems to be a minimum of 5 minutes and depends on the wattage of the microwave used, however there was an inverse relationship between the duration of microwave sterilization and the amount of viable bacterial colonies present on the catheter.^29^ Polyethylene catheters were shown to also be completely sterilized by a microwave sterilization method; however, this material of catheter was only evaluated in one study and no data regarding the physical properties of the catheter was present.^29^ Additionally, it has been determined that plastic catheters melted in the microwave.^28^ Microwave sterilization is also not a recommend cleaning method for other catheter material such as latex or PVC as there is evidence of melting and incomplete sterilization.^19,28^ Although gross changes in physical properties of catheters were documented in a few studies, less is known regarding the microscopic changes and changes in stiffness of catheters. It is crucial to acknowledge that based on expert consensus opinion, the physical properties of catheters are a crucial risk factor for ureteral micro trauma and development of UTIs.^17^ Other heat-based methods (steaming and boiling) were not effective and damaged PVC catheters.^19^

The quality of evidence for the efficacy of chemical cleaning is limited because the effectiveness of most have not been duplicated by more than one study, and many do not have corresponding data regarding their effects on catheter physical properties. To date, it seems that the most promising chemical cleaning solutions are 70% alcohol and Milton solution. It was demonstrated by one study that soaking a PVC catheter in the 70% alcohol solution for five minutes reduced bacterial colony counts by 6 log and did not affect the physical properties of PVC catheters. Soaking for longer period in alcohol solution is not recommended, as damage was seen in the catheters soaked for 45 minutes.^31^ Milton solution also did not affect the properties of PVC catheters, but did show some evidence of VNCB present.^19^ Other cleaning solutions such as 0.6% peroxide, a 1:4 solution of bleach with tap-water or a 1:2 solution of betadine in tap water were shown to be bactericidal against for *E. coli* but no structural analysis was performed.^34^ Another study supported the suggestion that peroxide could be a potential cleaning solution as it reported that a 3% peroxide solution reduced the bacterial colony count by 3 log; however, the same study also noted that it was less effective than rinsing with water and drying alone, which reduced the bacteria colony count by 5 log.^28^ Vinegar, while it appears to not affect the physical properties of the catheters, does not seem to be a suitable bactericidal cleaning solution.^19,28^ Kovindha et al. examined the effects of long term reuse with a Savlon cleaning solution as a case series by imaging catheters with EM.^7^ They found that there was encrustation on the lumen, and that the catheters had a 20% increase in stiffness. It should be noted that the two non-control catheters used in this case series were used for 1.5 and 2 years. Given the extent of time of reuse, one cannot make a conclusion on the short-term burden of reuse form this study. Future studies, such as randomized-controlled that incorporate larger sample sizes and long-term follow-up, should be conducted before these chemical cleaning solutions can be recommended for the reuse of catheters.

Mechanical cleaning methods such as washing with detergent or using a domestic jewelry cleaner left much to be desired when used on their own. When washing with detergent, one study reported that 44% of catheters still resulted in a positive culture^35^, while another showed that there was still 100,000 bacteria/mL present.^32^ A combined approach of washing with detergent and microwaving was better, but still resulted in 22% of catheters having a positive culture swab, and it was limited by the fact that microwaving greatly damages some catheter materials. A third study reported that just rinsing with tap water and drying alone resulted in a greater reduction in bacteria on catheters than washing with detergent alone.^28^ Just soaking in detergent also lowered counts of *E. coli* and *K. pneumonia* and did not damage PVC catheters, although it was reported to leave behind some residue in the lumen of the catheters.^16^ Soaking in detergent followed by soaking in Milton solution (“Milton method”) was reported as totally bactericidal and left no evidence of damage on PVC catheters.^19^ The other mechanical cleaning method analyzed, domestic jewelry cleaner, was not an effective as it was not bactericidal and caused surface damage to the catheters.^19^ Photocatalytic sterilization is an interesting proposal as it was reportedly bactericidal for all bacteria tested but no structural analysis was performed.^30^ A major limitation to this method is that a special TiO_2_ coated silicone catheter and ideally a black lamp is required, although it was reported that sunlight could be used as an alternative to the black lamp. In addition, we had to grade the study done by Sekiguchi et al. in terms of risk of bias as critical because there was missing data. 15 TiO_2_ catheters were analysed but only 10 control catheters analysed. There should have been the same number of control catheters as TiO_2_ catheters analyzed as there should be one of each per participant that completed the study hence there was missing data and therefore a risk of bias.

Newman et al. performed a cohort study where they did not control for the method of cleaning.^16^ 41% of participants stated they just rinsed with water and 41% reported they use both soap and water to clean their catheters before reuse. These catheters were collected, and it was found that 74% of them had microbial contamination and 20% had biofilm formation. Debris from residuals of urine, soap, detergent and cells were seen on EM. It should be noted that the mean number of days these catheters were reused prior to analysis was 21 days, but reuse ranged from one day to 270 days. Because of this wide range, there was some bias of generalizing the results of the impact of bacterial load.

### Limitations of this review

A major limitation of this systematic review was that every experimental study used a different approach for analyzing the effectiveness of cleaning methods. For example, the catheter material, method of cleaning, indicator organisms, and outcome measures were not constant among studies, making quality assessment and meaningful comparisons a challenge. The non-experimental studies also did not standardize the type of catheter or cleaning method used. Many of the studies also only performed the cleaning method once before analysis, which generated outcomes that were not practical for real-life application^36^ as those who reuse catheters often do so more than once. Many studies also did not completely analyze the impact of cleaning on the physical properties of the catheters, which also decreased the practicality of their outcomes.

The amount of published data on the topic is also a limitation on our results. While there were 12 studies that met our inclusion criteria, many of them were examining different cleaning methods on different catheter types, so in many cases cleaning methods were only examined in one study. For example, the two most promising cleaning methods reported, 70% alcohol immersion and “Milton method” were only examined in one study which decreased our confidence. As a result, we were not able to perform any meaningful meta-analysis.

## Conclusion

There were two cleaning methods reported that reduced all of the bacteria tested to negligible levels while confirming that the structural integrity of the catheter was maintained: soaking catheters in 70% ethanol solution for 5 minutes, and the combined approach of soaking catheters for 5 minutes in hot detergent water followed by soaking for 15 minutes in Milton solution. The structural integrity of the two cleaning methods were confirmed by calorimetric analysis and EM respectively. Major limitations to this result are that these methods of cleaning have not been confirmed effective by more than one study, as only PVC catheters were examined, and outcomes were measured after only a single reuse. While these cleaning methods are promising, additional data must be obtained before we would feel comfortable challenging current clinical recommendations. The authors believe that this topic requires further consideration and additional research that could provide better insight on cleaning methods, reuse of catheters, and potential benefits versus harm to individuals who use of catheters for IC.

## Data Availability

All available data is presented in this manuscript.

## Acknowledgments

Matthias Walter and Andrei V. Krassioukov had full access to all the data in the study and takes responsibility for the integrity of the data and the accuracy of the data analysis.

## Study concept and design

Mark Grasdal, Matthias Walter, and Andrei V. Krassioukov

## Acquisition of data

Mark Grasdal, Matthias Walter, and Andrei V. Krassioukov

## Analysis and interpretation of data

Mark Grasdal, Matthias Walter, and Andrei V. Krassioukov

## Drafting of the manuscript

Mark Grasdal

## Critical revision of the manuscript for important intellectual content

Matthias Walter and Andrei V. Krassioukov

## Statistical analysis

n/a

## Funding

Mark Grasdal (University of British Columbia – Faculty of Medicine Summer Student Research Program Award recipient) and Andrei V. Krassioukov (University of British Columbia – Faculty of Medicine, Department of Medicine, Endowed Chair in Rehabilitation Medicine).

### Administrative, technical, or material support

n/a

### Supervision

Matthias Walter and Andrei V. Krassioukov

### Other

N/A.

## References

1. Bakke, A., Irgens, L. M., Malt, U. F. & Hoisreter, P. A. (1993). Clean intermittent catheterisation performing abilities, aversive experiences and distress. Paraplegia. 31, 288–297.

2. Guttmann, L. & Frankel, H. (1966). The value of intermittent catheterisation in the early management of traumatic paraplegia and tetraplegia. Paraplegia. 4, 63–84.

3. Lapides, J., Diokno, A. C., Silber, S. M. & Lowe, B. S. (1972). Clean, intermittent self-catheterization in the treatment of urinary tract disease. J Urol. 107, 458–461.

4. Hill, T. C., Baverstock, R., Carlson, K., Estey, E.P., Gray, G.J., Hill, D.C., Ho, C., McGinnis, R.H., Moore, K., Parmar, R. (2013) Best practices for the treatment and prevention of urinary tract infection in the spinal cord injured population: The Alberta context Bladder management in the context of a spinal cord injury. Can Urol Assoc J. 77, 122–30.

5. Groen, J., Pannek, J., Castro Diaz, D., Del Popolo, G., Gross, T., Hamid, R., Karsenty, G., Kessler, T. M., Schneider, M., ‘T Hoen, L., Blok, B. (2016). Summary of European Association of Urology (EAU) Guidelines on Neuro-Urology. Eur Urol. 69, 324–333.

6. Wyndaele, J. J., Brauner, A., Geerlings, S. E., Bela, K., Peter, T., Bjerklund-Johanson, T. E., (2012). Clean intermittent catheterization and urinary tract infection: Review and guide for future research. BJU International. 110, E910–7.

7. Kovindha, A., Mai, W. N. C., Madersbacher, H. (2004). Reused silicone catheter for clean intermittent catheterization (CIC): Is it safe for spinal cord-injured (SCI) men? Spinal Cord. 42, 638–642.

8. Avery, M. Prieto P., Okamoto, I., Cullen, S., Clancy, B., Moore, K.N., Macaulay, M., Fader, M. (2018). Reuse of intermittent catheters: A qualitative study of IC users’ perspectives. BMJ Open. 8, 21554.

9. Prieto, J. A., Murphy, C., Moore, K. N., Fader, M (2014). WITHDRAWN: Intermittent Catheterisation for Long-Term Bladder Management. Cochrane Database Syst Rev. 2014 Sep 10;(9):CD006008. doi: 10.1002/14651858.CD006008.pub3. Update in: Cochrane Database Syst Rev. 2017 Aug 08;8:CD006008. PMID: 25208303.

10. Christison, K. Walter, M., Wyndaele, J.J.J.M., Kennelly, M., Kessler, T. M., Noonan V.K. Fallah, N., Krassioukov, A. (2018). Intermittent catheterization: The devil is in the details. J. Neurotrauma. 35, 985–989.

11. Gould, C. v., Umscheid, C. A., Agarwal, R. K., Kuntz, G., Pegues, D. A. (2010). Guideline for Prevention of Catheter-Associated Urinary Tract Infections 2009. Infection Control & Hospital Epidemiology. 31, 319–26.

12. Campeau, L. Shamout, S., Baverstock, R.J., Carlson, K.V., Elterman, D.S., Hickling, D.R., Steele, S.S., Welk, B. (2020). Canadian urological association best practice report: Catheter use. Can Urol Assoc J. 14, E281–E289.

13. Sun, A. J., Comiter, C. V., Elliott, C. S. (2018). The cost of a catheter: An environmental perspective on single use clean intermittent catheterization. Neurourol Urodyn. 37, 2204–2208.

14. Patel, D. N., Alabastro, C. G., Anger, J. T. (2018). Prevalence and Cost of Catheters to Manage Neurogenic Bladder. Current Bladder Dysfunction Reports. 13, 215–223.

15. Walter, M., Krassioukov, A. (2020). Single-use Versus Multi-use Catheters: Pro Single-use Catheters. Eur Urol Focus. 6, 807–808.

16. Newman, D. K., New, P.W., Heriseanu, R., Petronis, S., Håkansson, J., Håkansson, M. A., Lee, B. B., (2020). Intermittent catheterization with single-or multiple-reuse catheters: clinical study on safety and impact on quality of life. Int Urol Nephrol. 52, 1443–1451.

17. Kennelly, M., Thiruchelvam, N., Averbeck, M. A., Konstatinidis, C., Chartier-Kastler, E., Trøjgaard, P., Vaabengaard, R., Krassioukov, A., Jakobsen, B. P. (2019). Adult Neurogenic Lower Urinary Tract Dysfunction and Intermittent Catheterisation in a Community Setting: Risk Factors Model for Urinary Tract Infections. Adv Urol. 2757862.

18. Jeong, S. J., Oh, S. J. (2019). Recent updates in urinary catheter products for the neurogenic bladder patients with spinal cord injury. Korean J Neurotrauma. 15, 77–87.

19. Wilks, S. A., Morris, N. S., Thompson, R., Prieto, J. A., Macaulay, M., Moore, K. N., Keevil, C. W., Fader, M. (2020). An effective evidence-based cleaning method for the safe reuse of intermittent urinary catheters: In vitro testing. Neurourol Urodyn. 39, 907–915.

20. Håkansson, M. A. (2014). Reuse versus single-use catheters for intermittent catheterization: What is safe and preferred? Review of current status. Spinal Cord. 52, 511–516.

21. Sherbondy, A. L., Cooper, C. S., Kalinowski, S. E., Boyt, M. A., Hawtrey, C. E. (2002). Variability in catheter microwave sterilization techniques in a single clinic population. J Urol. 168, 562–564.

22. Liberati, A., Altman, D. G., Tetzlaff, J., Mulrow, C., Gøtzsche, P., Ioannidis, J. P. A., Clarke, M., Devereaux, P. J., Kleijnen, J., Moher, D. (2009) The PRISMA statement for reporting systematic reviews and meta-analyses of studies that evaluate health care interventions: Explanation and elaboration. PLoS Med. 6, e1000100.

23. Covidence systematic review software. Veritas Health Innovation: Melbourne, VIC, Australia [Available from: https://www.covidence.org] (Last accessed April, 2021).

24. Joanna Briggs Institute. “The Joanna Briggs Institute critical appraisal tools for use in JBI systematic reviews: checklist for quasi-experimental studies (non-randomized experimental studies) 2017 [Available from: https://jbi.global/critical-appraisal-tools] (Accessed April 2021).

25. National Heart, Lung, and Blood Institute. “Study quality assessment tools: quality assessment tool for observational cohort and cross-sectional studies.” [Available from: https://www.nhlbi.nih.gov/health-topics/study-quality-assessment-tools] (Accessed April 2021).

26. National Heart, Lung, and Blood Institute website. “Study Quality Assessment Tools: quality assessment tool for case series studies.” [Available from: https://www.nhlbi.nih.gov/health-topics/study-quality-assessment-tools] (Accessed April 2021).

27. Sterne, J. A. C, Hernán, M. A., Reeves, B. C., Savović, J., Berkman, N. D., Viswanathan, M., Henry, D., Altman, D. G., Ansari, M. T., Boutron, I., Carpenter, J. R. Chan, A-W., Churchill, R., Deeks, J. J., Hróbjartsson, A., Kirkham, J., Jüni, P., Loke, Y. K., Pigott, T. D., Ramsay, C. R., Regidor, D., Rothstein, H. R., Sandhu, L., Santaguida, P. L., Schünemann, H. J., Shea, B., Shrier, I., Tugwell, P., Turner, L., Valentine, J. C., Waddington, H., Waters, E., Wells, G. A., Whiting, P. F., Higgins, J. P. T. (2016). ROBINS-I: a tool for assessing risk of bias in non-randomised studies of interventions. BMJ. 355:i4919.

28. Lavallée, D. J., Lapierre, N. M., Henwood, P. K., Pivik, J. R., Best, M., Springthorpe, V. S., Sattar, S. A. (1995). Catheter cleaning for re-use in intermittent catheterization: new light on an old problem. SCI Nurs. 12, 10–12.

29. Griffith, D., Nacey, J., Robinson, R., Delahunt, B. (1993). Microwave Sterilization of Polyethylene Catheters for Intermittent Self-Catheterization. Aust N Z J Surg. 63, 203–204.

30. Sekiguchi, Y., Yao, Y., Ohko, Y., Tanaka, K., Ishido, T., Fujishima, A., Kubota, Y. (2007). Self-sterilizing catheters with titanium dioxide photocatalyst thin films for clean intermittent catheterization: Basis and study of clinical use. Int J Urol. 14, 426–430.

31. Bogaert, G. A. Goeman, L., Ridder, D. D., Wevers, M., Ivens, J., Schuermans, (2004). The physical and antimicrobial effects of microwave heating and alcohol immersion on catheters that are reused for clean intermittent catheterisation. Eur Urol. 46, 641–646.

32. Mervine, J., Temple, R. (1997). Using a microwave oven to disinfect intermittent-use catheters. Rehabil Nurs. 22, 318–320.

33. Douglas, C., Burke, B., Kessler, D. L., Cicmanec, J. F., Bracken, B. (1990). Method for Sterilizing Urinary Catheters. Urology. 35, 219–222.

34. Kurtz, M. J., van Zandt, K., Burns, J. L. (1995). Comparison Study of Home Catheter Cleaning Methods. Rehabil Nurs. 20, 212–214.

35. Chan, J. L., Cooney, T. E., Schober, J. M. (2009). Adequacy of sanitization and storage of catheters for intermittent use after washing and microwave sterilization. J Urol. 182, 2085–2089.

36. Krassioukov, A., Cragg, J. J., West, C., Voss, C., Krassioukov-Enns, D. (2015). The good, the bad and the ugly of catheterization practices among elite athletes with spinal cord injury: A global perspective. Spinal Cord. 53, 78–82.

