## Supplementary Appendix SA1 for "The microbiological and physical properties of catheters for intermittent catheterization: A systematic review on the impact of reuse and cleaning methods"

**Supplementary Material Appendix 1: Search Strategy**

**MEDLINE (Ovid) May 13, 2020**

Database: Ovid MEDLINE(R) and Epub Ahead of Print, In-Process & Other Non-Indexed Citations, Daily and Versions(R) <1990 to May 13, 2020>

Search Strategy:

--------------------------------------------------------------------------------

1     ("intermittent catheter*" or "urinary catheter*" or "bladder catheter*").mp. [mp=title, abstract, heading word, drug trade name, original title, device manufacturer, drug manufacturer, device trade name, keyword, floating subheading word, candidate term word]

2     ("clean*" or "cleaning methods" or "cleaning solution" or "disinfect" or "decontaminate" or "steril*").mp. [mp=title, abstract, heading word, drug trade name, original title, device manufacturer, drug manufacturer, device trade name, keyword, floating subheading word, candidate term word]

3     ("physical" or "mechanical" or "microscopic").mp. [mp=title, abstract, heading word, drug trade name, original title, device manufacturer, drug manufacturer, device trade name, keyword, floating subheading word, candidate term word]

4     ("re-use" or "reuse*").mp. [mp=title, abstract, heading word, drug trade name, original title, device manufacturer, drug manufacturer, device trade name, keyword, floating subheading word, candidate term word]

5     2 or 3 or 4

6     1 and 5

7     limit 6 to yr="1990 -Current"

**Hits: 2375**

**Embase (Ovid) May 14, 2020**

Database: Embase <1990 to May 14, 2020>

Search Strategy:

--------------------------------------------------------------------------------

1	("intermittent catheter*" or "urinary catheter*" or "bladder catheter*") .mp. [mp=title, abstract, heading word, drug trade name, original title, device manufacturer, drug manufacturer, device trade name, keyword, floating subheading word, candidate term word]

2	("clean*" or "cleaning methods" or "cleaning solution" or "disinfect" or "decontaminate" or "steril*").mp. [mp=title, abstract, heading word, drug trade name, original title, device manufacturer, drug manufacturer, device trade name, keyword, floating subheading word, candidate term word]

3	("physical" or "mechanical" or "microscopic").mp. [mp=title, abstract, heading word, drug trade name, original title, device manufacturer, drug manufacturer, device trade name, keyword, floating subheading word, candidate term word]

4	("re-use" or "reuse*").mp. [mp=title, abstract, heading word, drug trade name, original title, device manufacturer, drug manufacturer, device trade name, keyword, floating subheading word, candidate term word]

5	2 or 3 or 4

6	1 and 5

7	limit 6 to yr="1990 -Current"

**Hits: 4259**

**CINAHL Complete May 20, 2020**

Database: CINAHL Complete <1990 to May 20, 2020>

Interface - EBSCOhost Research Databases

S1	"intermittent catheter*" or "urinary catheter*" or "bladder catheter*"	Search modes - Boolean/Phrase	Interface - EBSCOhost Research Databases

S2	"clean*" or "cleaning methods" or "cleaning solution" or "disinfect" or "decontaminate" or "steril*"	Search modes - Boolean/Phrase

S3	"physical" or "mechanical" or "microscopic"	Search modes - Boolean/Phrase

S4	"re-use" or "reuse*"	Search modes - Boolean/Phrase

S5	S2 OR S3 OR S4	Search modes - Boolean/Phrase

S6	S1 AND S5	Search modes - Boolean/Phrase

S7	S1 AND S5	Limiters - Published Date: 19900101-20201231

**Hits: 835**

**Web of Science Core Collection May 20, 2020**

Indexes=SCI-EXPANDED, SSCI, A&HCI, CPCI-S, CPCI-SSH, ESCI Timespan=1990-2020

1	ALL=("intermittent catheter*" or "urinary catheter*" or "bladder catheter*")

2	ALL=("clean*" or "cleaning methods" or "cleaning solution" or "disinfect" or "decontaminate" or "steril*") (486,693)

3	ALL=("physical" or "mechanical" or "microscopic")

4	ALL = ("re-use" or "reuse*")

5	#4 OR #3 OR #2

6	#5 AND #1

**Hits: 1971**

**EBM Reviews - Cochrane Central Register of Controlled Trials May 20, 2020**

1	("intermittent catheter*" or "urinary catheter*" or "bladder catheter*").mp. [mp=title, original title, abstract, mesh headings, heading words, keyword]

2	("clean*" or "cleaning methods" or "cleaning solution" or "disinfect" or "decontaminate" or "steril*").mp. [mp=title, original title, abstract, mesh headings, heading words, keyword]

3	("physical" or "mechanical" or "microscopic").mp. [mp=title, original title, abstract, mesh headings, heading words, keyword]

4	("re-use" or "reuse*").mp. [mp=title, original title, abstract, mesh headings, heading words, keyword]

5	2 or 3 or 4

6	1 and 5

7	limit 6 to yr="1990 -current”

**Hits:** **173**
