## Supplementary Appendix SA2 for "The microbiological and physical properties of catheters for intermittent catheterization: A systematic review on the impact of reuse and cleaning methods"

**Supplementary Material Appendix 2: Critical Appraisal Checklists**

**Table 1. JBI Critical Appraisal Checklist for Quasi-Experimental Studies (non-randomized experimental studies)**

| **Checklist** | Wilks et al., 2020 | Chan et al. 2009 | Sekiguchi et al. 2007 | Bogaert et al. 2004 | Mervine et al. 1997 | Lavallee et al. 1995 | Kurtz et al. 1995 | Griffith et al. 1993 | Douglas et al. 1990 |
| --- | --- | --- | --- | --- | --- | --- | --- | --- | --- |
| Is it clear in the study what is the ‘cause’ and what is the ‘effect’ | Yes | Yes | Yes | Yes | Yes | Yes | Yes | Yes | Yes |
| Were the participants included in any comparisons similar? | Yes (but catheters, not participants) | Yes (but catheters, not participants) | Yes (but catheters, not participants) | Yes (but catheters, not participants) | Yes (but catheters, not participants) | Yes (but catheters, not participants) | Yes (but catheters, not participants) | Yes (but catheters, not participants) | Yes (but catheters, not participants) |
| Were the participants included in any comparisons receiving similar treatment/ care, other than the exposure or intervention of interest? | Not applicable | Not applicable | Not applicable | Not Applicable | Not Applicable | Not applicable | Not applicable | Not Applicable | Not Applicable |
| Was there a control group? | Yes | Yes | Yes | Yes | No | Yes | Yes | Yes | Yes |
| Were there multiple measurements of the outcome both pre and post the intervention/exposure? | No | No | Yes | No | No | No | No | No | No |
| Was follow up complete and if not, were differences between  groups in terms of their follow up adequately described and  analyzed? | Not Applicable | Not Applicable | Not applicable | Not applicable | Not applicable | Not Applicable | Not applicable | Not applicable | Not applicable |
| Were the outcomes of participants included in any comparisons measured in the same way? | Yes (but catheters, not participants) | Yes (but catheters, not participants) | Yes (but catheters, not participants) | Yes (but catheters, not participants) | Yes (but catheters, not participants) | Yes (but catheters, not participants) | Yes (but catheters, not participants) | Yes (but catheters, not participants) | Yes (but catheters, not participants) |
| Were outcomes measured in a reliable way? | Yes | Yes | Yes | Yes | Yes | Yes | Yes | Yes | Yes |
| Was appropriate statistical analysis used? | No | Yes | No | No | No | Yes | Yes | No | No |
| **Overall Appraisal** | **Include** | **Include** | **Include** | **Include** | **Include** | **Include** | **Include** | **Include** | **Include** |

**Table 2. NIH Quality Assessment Tool for Observational Cohort and Cross-Sectional Studies**

| **Criteria** | Newman et al., 2020 | Sekiguchi et al., 2007 | Sherbondy et al.,2002 |
| --- | --- | --- | --- |
| 1. Was the research question or objective in this paper clearly stated? | Yes | Yes | Yes |
| 2. Was the study population clearly specified and defined? | Yes | Yes | Yes |
| 3. Was the participation rate of eligible persons at least 50%? | Yes | Yes | Yes |
| 4. Were all the subjects selected or recruited from the same or similar populations (including the same time period)? Were inclusion and exclusion criteria for being in the study prespecified and applied uniformly to all participants? | Yes | Yes | Yes |
| 5. Was a sample size justification, power description, or variance and effect estimates provided? | Yes | No | No |
| 6. For the analyses in this paper, were the exposure(s) of interest measured prior to the outcome(s) being measured? | Yes | Yes | Yes |
| 7. Was the timeframe sufficient so that one could reasonably expect to see an association between exposure and outcome if it existed? | Yes | Yes | Yes |
| 8. For exposures that can vary in amount or level, did the study examine different levels of the exposure as related to the outcome (e.g., categories of exposure, or exposure measured as continuous variable)? | No | No | No |
| 9. Were the exposure measures (independent variables) clearly defined, valid, reliable, and implemented consistently across all study participants? | No | Yes | Yes |
| 10. Was the exposure(s) assessed more than once over time? | No | No | No |
| 11. Were the outcome measures (dependent variables) clearly defined, valid, reliable, and implemented consistently across all study participants? | Yes | Yes | Yes |
| 12. Were the outcome assessors blinded to the exposure status of participants? | Not applicable | Not applicable | Not applicable |
| 13. Was loss to follow-up after baseline 20% or less? | Yes | Yes | No |
| 14. Were key potential confounding variables measured and adjusted statistically for their impact on the relationship between exposure(s) and outcome(s)? | Yes | No | No |
| **Quality Rating** | **Fair** | **Fair** | **Fair** |

**Table 3. NIH Tool for Case Series Studies**

| **Criteria** | Kovindha et al., 2004 |
| --- | --- |
| Was the study question or objective clearly stated? | Yes |
| Was the study population clearly and fully described, including a case definition? | Yes |
| Were the cases consecutive? | Yes |
| Were the subjects comparable? | Yes |
| Was the intervention clearly described? | Yes |
| Were the outcome measures clearly defined, valid, reliable, and implemented consistently across all study participants? | Yes |
| Was the length of follow-up adequate? | Not applicable |
| Were the statistical methods well-described? | Not applicable |
| Were the results well-described? | Yes |
| **Quality Rating** | **Good** |
